# Geographic heterogeneity of gut microbiome biomarkers across neurological conditions: sub-national discordance in ten cohorts from eleven cities in six countries

**DOI:** 10.64898/2026.11.19.26368539

**Authors:** Johann Tello Meryk

## Abstract

Gut microbiota composition has been associated with multiple neurological conditions, and candidate biomarkers have been proposed on the basis of these associations. Their usefulness presupposes that taxa altered in one population are also altered in others, a condition that is rarely verified. This work quantifies the extent to which it holds, and the geographic scale at which it ceases to do so.

Ten public 16S rRNA gene sequencing cohorts of Alzheimer’s disease, Parkinson’s disease and autism spectrum disorder, collected in eleven cities across six countries, were reprocessed from raw data, comprising 1,185 samples with uniform processing within each condition.

Models trained and evaluated within a single cohort reached a mean AUC of 0.751 in Alzheimer’s disease, 0.741 in autism spectrum disorder and 0.673 in Parkinson’s disease. When applied to a different population, these values fell to 0.569, 0.542 and 0.551 respectively. The decline was not associated with geographic distance: transfer between cities within the same country was equivalent to transfer between continents. No taxon appeared among the twenty most important features in all cohorts of a given condition, and no correlation between rankings reached significance.

Classifiers trained to predict cohort of origin, disregarding clinical group, reached AUC values of 0.87 to 1.00 using control subjects, and equivalent values using affected subjects. In one cohort with two recruitment centers and a single protocol, controls from both cities were separable (AUC 0.947), models did not transfer between centers (0.562 and 0.515), and their rankings correlated 0.003.

Geographic heterogeneity of discriminant taxa operates below the national scale, which constrains the generalization of microbiome signatures derived from individual cohorts.

## Introduction

The human gut microbiota harbors hundreds of bacterial genera whose metabolic activity influences host physiology beyond the digestive tract. Over the past two decades, a set of bidirectional communication pathways between the gut and the central nervous system has been characterized, collectively known as the gut-brain axis [1]. Among the described pathways are bacterial production of short-chain fatty acids, which modulate intestinal barrier integrity and microglial activity [2]; tryptophan metabolism toward indolic derivatives and the kynurenine pathway, some of whose products cross the blood-brain barrier [3]; afferent signaling through the vagus nerve [4]; and modulation of systemic inflammation by bacterial wall components [1]. These pathways provide a plausible mechanism by which alterations in microbiota composition might be associated with neurological, neurodegenerative and neurodevelopmental conditions.

On this basis, a considerable body of microbial composition studies has accumulated in patients with Alzheimer’s disease [5,6,7,9], Parkinson’s disease [8] and autism spectrum disorder [10,11,12]. The predominant design is case-control, with 16S rRNA gene amplicon sequencing and differential abundance analysis between groups. These studies consistently identify taxa whose abundance differs between subjects with the condition and controls from the same population; several propose panels of candidate biomarkers, and some train classifiers that achieve appreciable performance within their own cohort [8,13]. Throughout this work, “discriminant taxon” is used to refer to a genus whose abundance differs between groups, and “candidate biomarker” when referring to taxa proposed as such in the prior literature; neither term implies diagnostic validation.

Aggregating these findings in reviews and meta-analyses presupposes that taxa altered in one population are also altered in others. This assumption has rarely been tested directly, because doing so requires reprocessing other investigators’ cohorts from raw data under uniform criteria.

The dominant expectation in the field takes two forms. The more widespread presupposes that taxa altered in a condition are altered independently of population, and candidate biomarker panels have been proposed on that basis. A second form, present in studies that acknowledge geographic variation in the microbiota, admits differences between regions or countries but implicitly maintains comparability within a single territory. Neither anticipates discordance between cities of the same country, still less between recruitment centers within a single study.

Regional variation in the gut microbiota has already shown consequences for model generalization. In a cohort of 7,009 individuals from fourteen districts of a single Chinese province, host location was the factor most strongly associated with microbial variation, and metabolic disease models derived in one locality failed when applied in another [14]. It remains unknown, however, whether this phenomenon extends to neurological and neurodevelopmental conditions, whether it also operates at international scale, and whether it affects the identity of discriminant taxa in addition to aggregate model performance. A second question overlaps with this one: comparison between independent studies accumulates differences in DNA extraction, library preparation, amplified hypervariable region, platform and sequencing depth [15]. These differences are well known, but verifying them beforehand is not part of standard practice when aggregating cohorts.

External validation in an independent population is, moreover, an established requirement for predictive models in health, set out in current reporting guidelines [21,22]. In the microbiome literature, however, classification models are usually reported with internal validation only, and the effect of the derivation population on their performance is not quantified systematically.

This work addresses both questions through uniform reprocessing of ten public 16S sequencing cohorts, corresponding to three conditions, collected in eleven cities across six countries. We evaluated, first, the capacity to separate cases from controls within each cohort; second, the transferability of that separation when the model is applied to a different population; and third, the concordance between the taxa that sustain the separation in each population. To distinguish the geographic component from the technical one, we separately analyzed a cohort with two recruitment centers in different cities, processed under a single protocol.

## Results

### Cohort selection and provenance

Searching public repositories identified approximately thirty deposits with 16S sequencing data in the three conditions. An admission protocol was applied to the public metadata before downloading any file, verifying amplified hypervariable region, sequencing platform, sample type, one-subject-per-sample uniqueness and availability of group labels.

Twenty deposits did not meet the criteria. Fourteen were excluded for technical incompatibility: 16S region different from the rest of the condition (n = 4), shotgun sequencing or RNA-Seq (n = 4), obsolete platform or proprietary format (n = 2), no accessible raw data (n = 2), and organism or gene different from that declared (n = 2). Six were excluded for design characteristics: therapeutic intervention or multiple time points per subject, pseudoreplication, and absence of group labels.

Ten cohorts met the criteria: five of Alzheimer’s disease, three of autism spectrum disorder and two of Parkinson’s disease, totaling 1,185 samples collected in eleven cities across six countries (Figure 1, Table 1).

**Figure 1.**
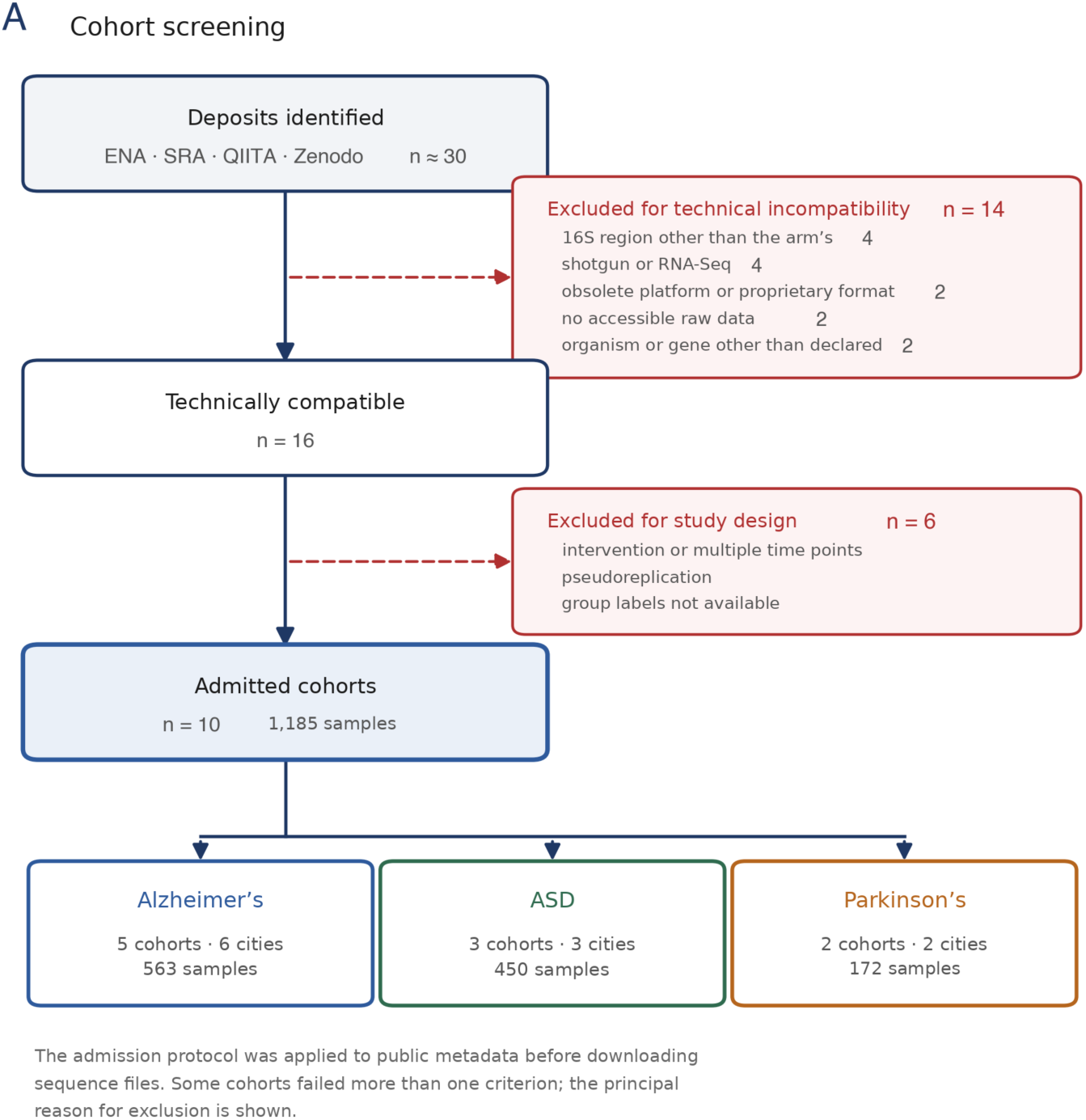
A. Cohort screening. Deposits identified in public repositories and reasons for exclusion. The admission protocol was applied to public metadata before downloading sequence files. Some cohorts failed more than one criterion; the principal reason for exclusion is shown.

**Figure 1.**
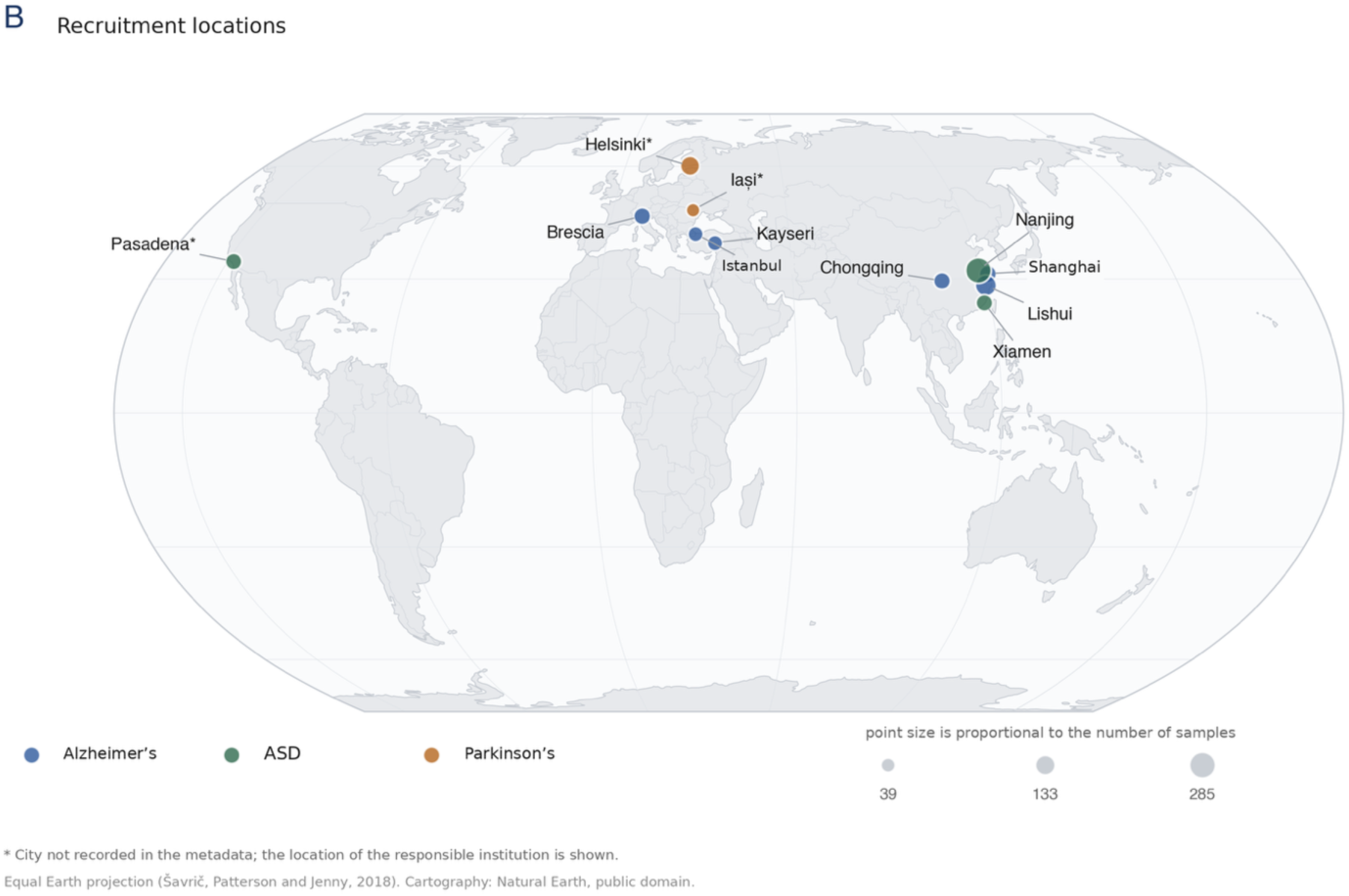
B. Recruitment locations. Point diameter is proportional to the number of samples. Asterisks mark the three cohorts whose recruitment city is not recorded in the metadata; the location of the responsible institution is shown instead. Equal Earth projection (Šavrič, Patterson and Jenny, 2018). Cartography: Natural Earth, public domain.

**Table 1.**
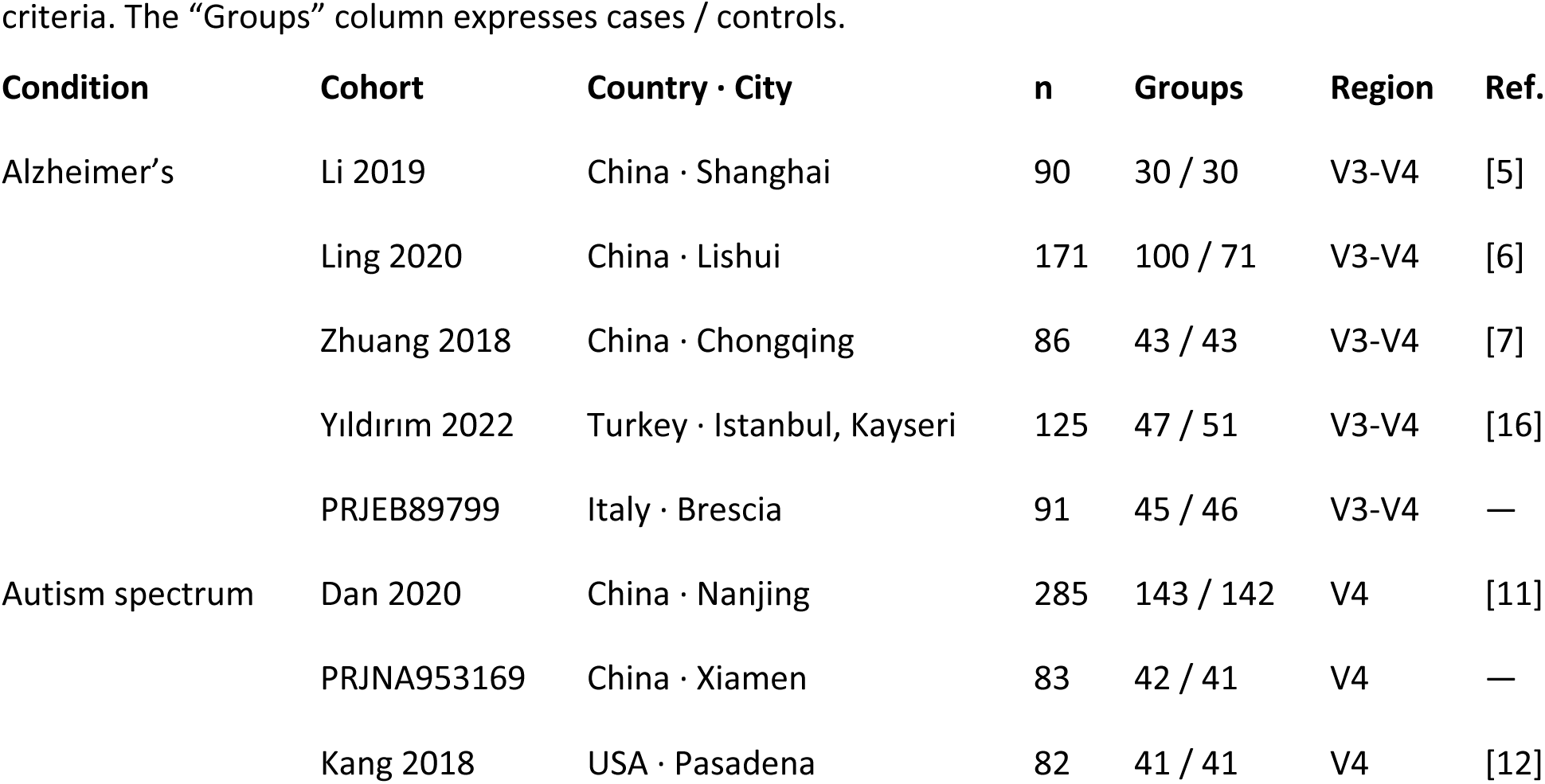
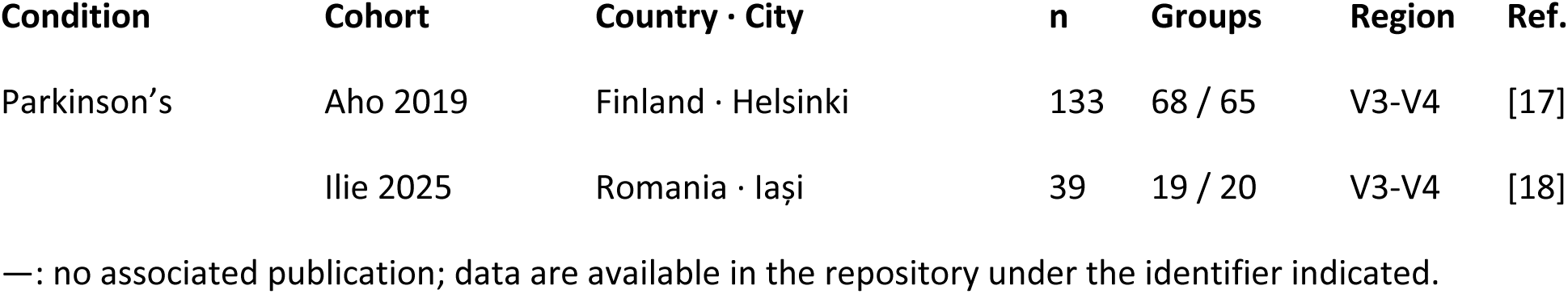
Admitted cohorts. n indicates the number of samples analyzed after applying the selection criteria. The “Groups” column expresses cases / controls.

| **Condition** | **Cohort** | **Country · City** | **n** | **Groups** | **Region** | **Ref.** |
|---|---|---|---|---|---|---|
| Alzheimer’s | Li 2019 | China · Shanghai | 90 | 30 / 30 | V3-V4 | [5] |
|  | Ling 2020 | China · Lishui | 171 | 100 / 71 | V3-V4 | [6] |
|  | Zhuang 2018 | China · Chongqing | 86 | 43 / 43 | V3-V4 | [7] |
|  | Yıldırım 2022 | Turkey · Istanbul, Kayseri | 125 | 47 / 51 | V3-V4 | [16] |
|  | PRJEB89799 | Italy · Brescia | 91 | 45 / 46 | V3-V4 | — |
| Autism spectrum | Dan 2020 | China · Nanjing | 285 | 143 / 142 | V4 | [11] |
|  | PRJNA953169 | China · Xiamen | 83 | 42 / 41 | V4 | — |
|  | Kang 2018 | USA · Pasadena | 82 | 41 / 41 | V4 | [12] |
| Parkinson’s | Aho 2019 | Finland · Helsinki | 133 | 68 / 65 | V3-V4 | [17] |
|  | Ilie 2025 | Romania · Iași | 39 | 19 / 20 | V3-V4 | [18] |

—: no associated publication; data are available in the repository under the identifier indicated.

### Processing reproduces published effect directions

To verify that uniform reprocessing did not introduce systematic deviations, the effect directions obtained were compared with those reported by the original authors in three Alzheimer’s disease cohorts [5,6,7]. Of fifteen evaluable comparisons, fourteen agreed in direction, thirteen of them with statistical significance in the reanalysis. The remaining discrepancy corresponded to an annotation error in extracting the published direction, not to divergence in processing. Internal performance obtained in the Turkish cohort (AUC 0.633) likewise matched that published by its authors (0.63) [16].

### Separation between cases and controls within each cohort

Models trained and evaluated within a single cohort reached AUC values between 0.583 and 0.955 (Figure 2). The mean was 0.751 in Alzheimer’s disease, 0.741 in autism spectrum disorder and 0.673 in Parkinson’s disease.

**Figure 2.**
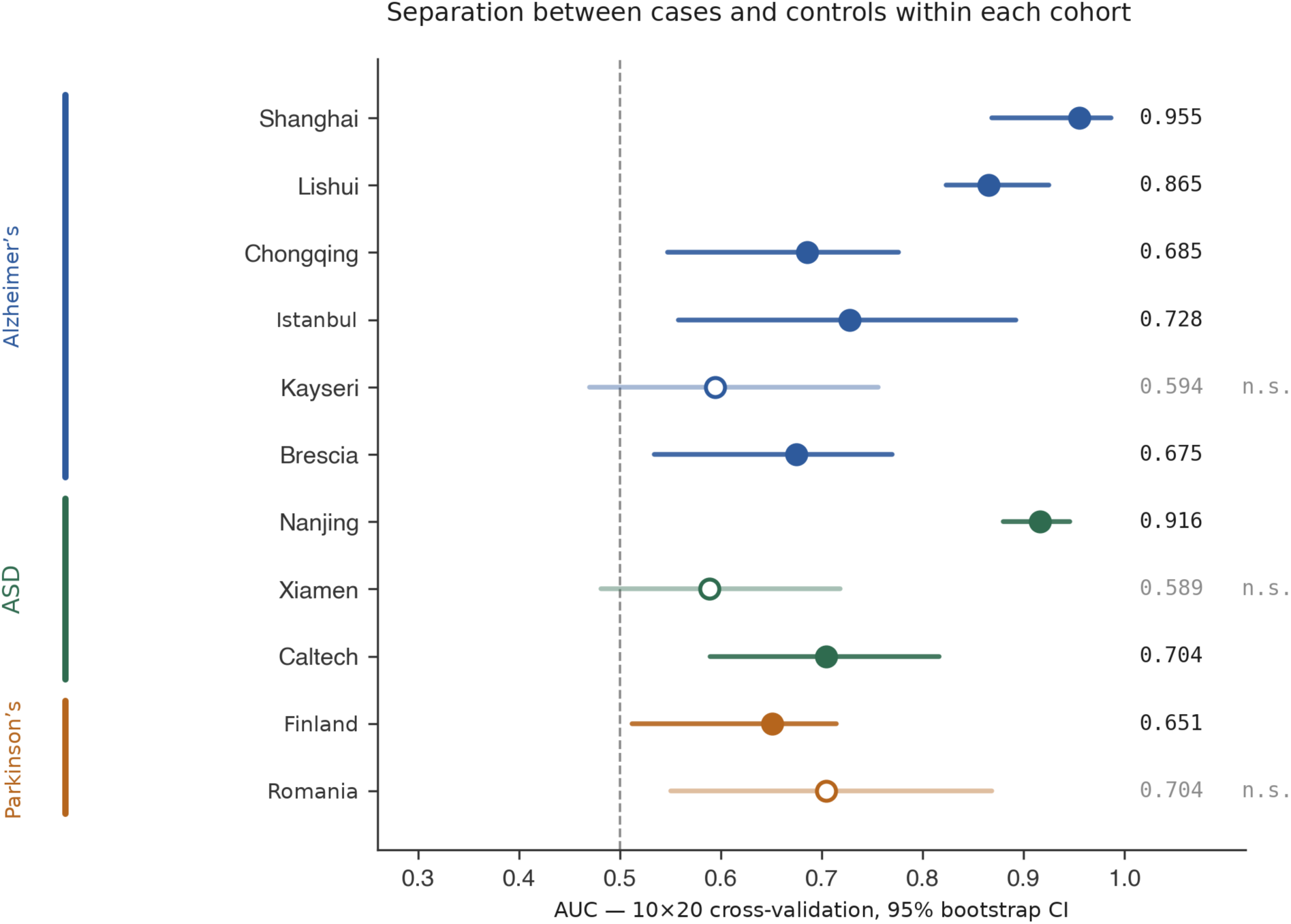
Separation between cases and controls within each cohort. AUC obtained by stratified ten-fold cross-validation, repeated twenty times with independent seeds. Horizontal bars represent 95% confidence intervals from 1,000 bootstrap resamples. Open points mark cohorts that did not reach significance in the permutation test (n.s.). The dashed line marks chance-level performance.

Three cohorts did not reach significance in the permutation test: Kayseri (AUC 0.594; p = 0.233), Xiamen (0.589; p = 0.156) and Iași (0.704; p = 0.053). All three correspond to the smallest sample sizes in the set. Three cohorts showed, by contrast, values above the range usually reported in the literature for these conditions: Shanghai (0.955), Lishui (0.865) and Nanjing (0.916). In two of them, a significant difference in sequencing depth was detected between cases and controls (Shanghai p = 0.007; Lishui p = 0.003), consistent with processing in separate batches by group.

In the case of Xiamen, a sensitivity analysis showed that internal performance depends on processing mode: when the cohort is reprocessed in paired-end mode, the AUC rises to 0.703 (p = 0.003). The reported absence of significance therefore corresponds to the uniform processing adopted for the condition, not to an absence of signal in the cohort.

### Separation does not transfer between populations

When each model was applied to a different cohort, the AUC declined in all three conditions. The mean across cross-cohort pairs was 0.569 in Alzheimer’s disease (30 pairs), 0.542 in autism spectrum disorder (6 pairs) and 0.551 in Parkinson’s disease (2 pairs), compared with internal values of 0.751, 0.741 and 0.673 respectively (Figure 3). In the Alzheimer’s disease condition, nine of thirty pairs fell below 0.5.

**Figure 3.**
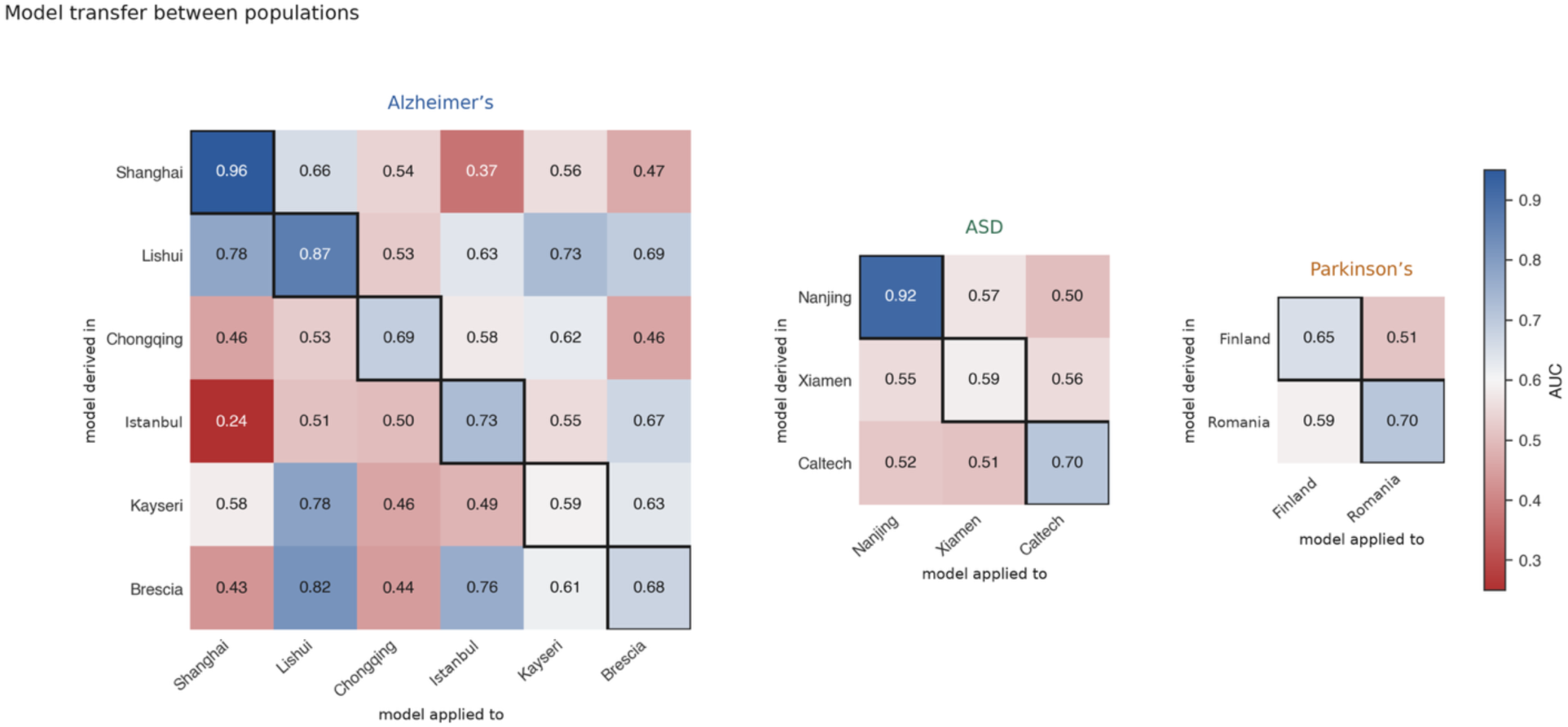
Model transfer between populations. Each cell shows the AUC obtained by training the model on the whole cohort indicated in the row and evaluating it on the whole cohort indicated in the column, restricted to shared features. The boxed diagonal corresponds to internal performance.

The magnitude of the decline was not associated with geographic distance. In Alzheimer’s disease, the mean AUC between cities of the same country was 0.565, and between different countries 0.571. In autism spectrum disorder, within-country transfer (0.563) likewise did not exceed between-country transfer (0.523).

### The taxa that sustain the separation do not coincide

Before comparing across cohorts, the stability of importance rankings within each cohort was evaluated. Spearman correlation between twenty independent fits with different seeds ranged from 0.896 to 0.959, indicating that estimation variability is low and that the discordance observed between cohorts is not attributable to fitting noise.

Across cohorts, no ranking correlation reached significance in any of the three conditions (Figure 4). Overlap among the twenty most important taxa ranged from 0 to 7 of 20 depending on the pair. No taxon appeared among the top twenty of all cohorts of Alzheimer’s disease or of autism spectrum disorder.

**Figure 4.**
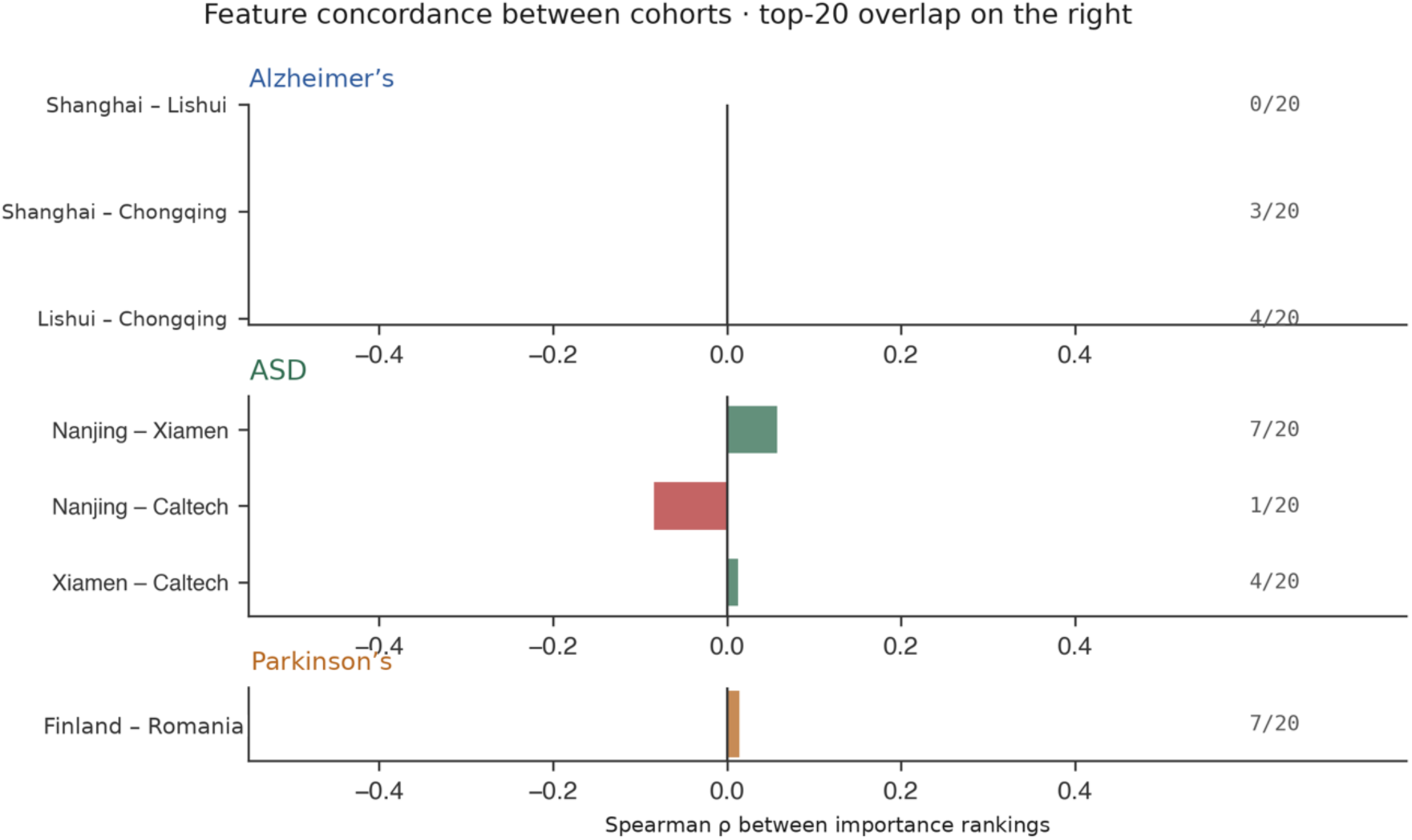
Feature concordance between cohorts. Spearman correlation between the averaged importance rankings of each pair of cohorts. The figure to the right of each bar indicates the number of taxa shared among the twenty most important in both cohorts of the pair.

The region-grouped contrast identified few taxa with concordant direction and significance: ten of 211 genera in Alzheimer’s disease, two of 178 in autism spectrum disorder and eight of 173 in Parkinson’s disease (Table 2). Most correspond to short-chain fatty acid producers. Three, nine and nine confirmed direction reversals were identified, respectively (Figure 5).

**Figure 5.**
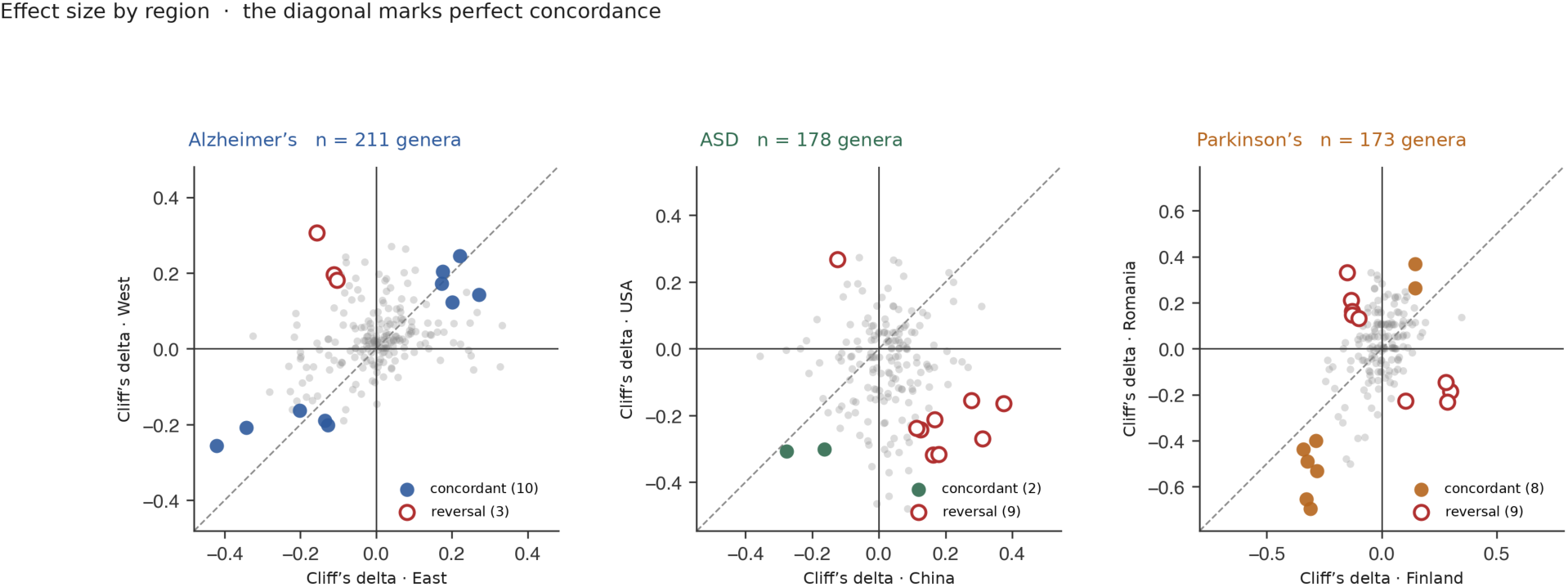
Effect size by region. Each point represents a genus. The axes show effect size (Cliff’s delta, cases versus controls) calculated separately in each region. Solid points mark concordant taxa: same sign, magnitude equal to or greater than 0.10 in both regions, and significance in both; open points mark confirmed reversals with non-overlapping confidence intervals. The diagonal marks perfect concordance.

**Table 2.** Taxa concordant across regions. A genus was considered concordant if it had the same sign in both regions, magnitude equal to or greater than 0.10 in both, and statistical significance in both. δ is Cliff’s delta (cases versus controls), calculated separately in each region; p values are from the Mann-Whitney U test. The columns “δ region 1” and “δ region 2” correspond, in that order, to the two regions indicated in parentheses in the first column: East and West for Alzheimer’s disease, China and USA for autism spectrum disorder, and Finland and Romania for Parkinson’s disease.

| Condition | Genus | δ region 1 | p | δ region 2 | p |
|---|---|---|---|---|---|
| Alzheimer’s | *Lachnospira* | -0.422 | <0.0001 | -0.256 | 0.002 |
| (East / West) | *Faecalibacterium* | -0.343 | <0.0001 | -0.208 | 0.014 |
|  | *Eisenbergiella* | +0.271 | <0.0001 | +0.143 | 0.019 |
|  | *Ruthenibacterium* | +0.221 | 0.0006 | +0.245 | 0.003 |
|  | *Haemophilus* | -0.202 | <0.0001 | -0.163 | 0.022 |
|  | Hungatella | +0.201 | 0.0003 | +0.123 | 0.040 |
|  | *Anaerotruncus* | +0.175 | 0.0005 | +0.204 | 0.0007 |
|  | *Collinsella* | +0.173 | 0.007 | +0.172 | 0.029 |
|  | *Clostridium* | -0.136 | 0.034 | -0.189 | 0.023 |
|  | *Fusicatenibacter* | -0.128 | 0.044 | -0.201 | 0.017 |
| Autism spectrum | *Segatella* | -0.277 | <0.0001 | -0.308 | 0.013 |
| (China / USA) | *Mediterraneibacter* | -0.163 | 0.007 | -0.302 | 0.019 |
| Parkinson’s | *Fusicatenibacter* | -0.310 | 0.002 | -0.695 | 0.0002 |
| (Finland / Romania) | *Blautia* | -0.328 | 0.001 | -0.653 | 0.0005 |
|  | *Roseburia* | -0.281 | 0.005 | -0.532 | 0.004 |
|  | *Agathobaculum* | -0.323 | 0.001 | -0.490 | 0.005 |
|  | *Agathobacter* | -0.340 | 0.0007 | -0.437 | 0.020 |
|  | *Erysipelotrichaceae* UCG-003 | -0.287 | 0.004 | -0.400 | 0.032 |
|  | *Ligilactobacillus* | +0.145 | 0.004 | +0.368 | 0.036 |
|  | *Porphyromonas* | +0.144 | 0.016 | +0.263 | 0.017 |

**Table 3.** Between-cohort separability by condition. AUC of classifiers trained to predict cohort of origin, disregarding clinical group. The range covers all cohort pairs of each condition.

| **Condition** | **Pairs** | **All subjects** | **Controls only** | **Cases only** |
|---|---|---|---|---|
| Alzheimer’s | 15 | 0.929 – 0.986 | 0.867 – 1.000 | 0.901 – 0.993 |
| Autism spectrum | 3 | 0.998 – 1.000 | 0.989 – 1.000 | 0.996 – 1.000 |
| Parkinson’s | 1 | 1.000 | 0.996 | 0.999 |

Parkinson’s disease illustrates that the two properties are independent: eight genera maintained concordant direction and significance between Finland and Romania, the highest directional concordance of the three conditions; nevertheless, the model trained in Finland performed at chance level in Romania (AUC 0.513) and the correlation between rankings was 0.014.

### Sample origin predominates over condition

To gauge the magnitude of variation between populations, classifiers were trained to predict the cohort of origin of each sample, disregarding clinical group. Using control subjects only, cohorts were separable with AUC values between 0.867 and 1.000. Using only subjects with the condition, values were equivalent: 0.942 on average in Alzheimer’s disease, 0.999 in autism spectrum disorder and 0.999 in Parkinson’s disease, with differences relative to controls of −0.026, +0.002 and +0.004 (Figure 6).

**Figure 6.**
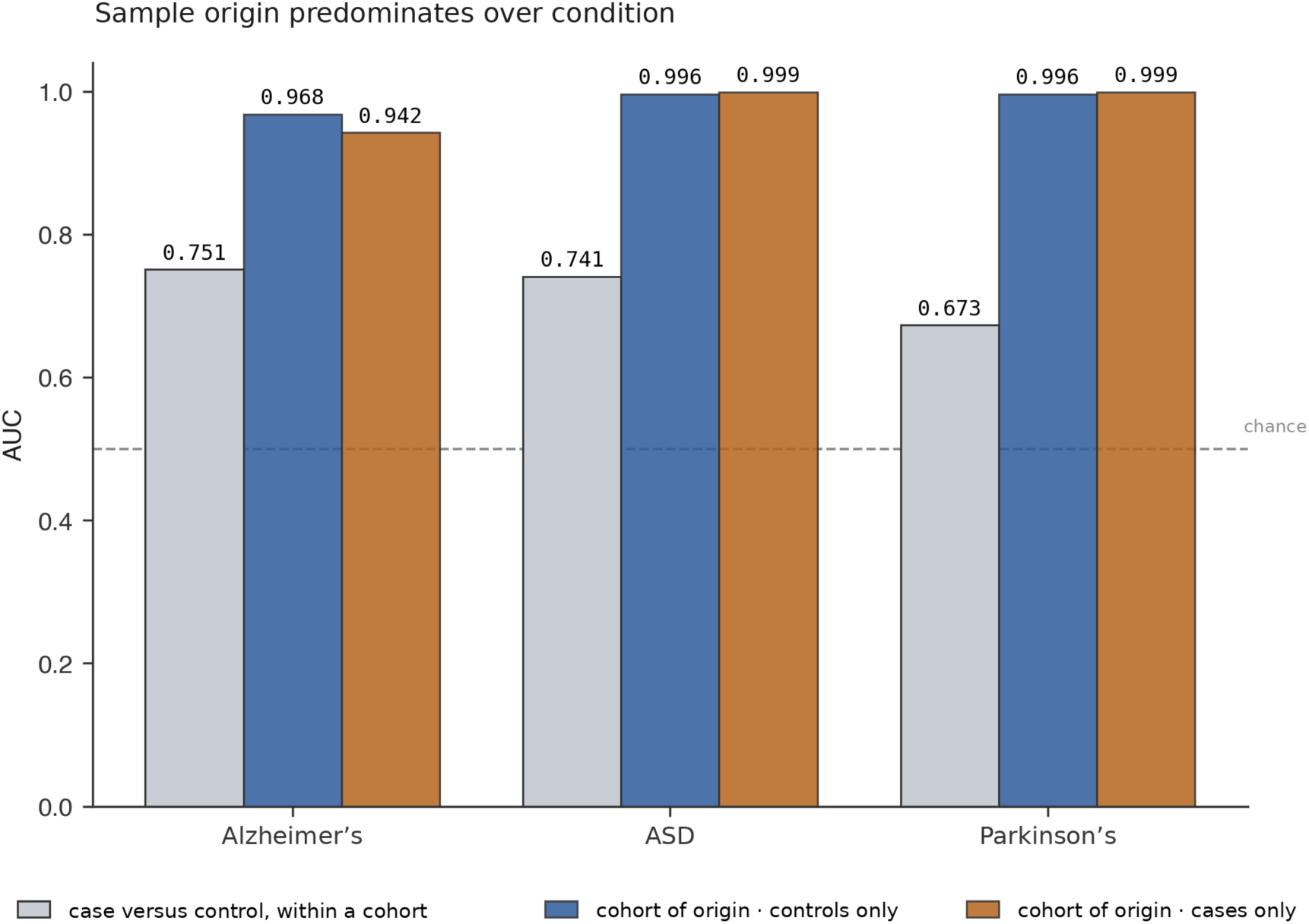
Sample origin predominates over condition. Gray bars: AUC for separating cases from controls within a single cohort, averaged over the cohorts of each condition. Blue and orange bars: AUC for separating cohorts when predicting sample provenance, using only control subjects or only subjects with the condition, respectively. The dashed line marks chance-level performance.

Determining which cohort a sample came from was therefore considerably easier than determining whether the subject had the condition, and this held in both healthy and affected subjects.

### Geographic separation under a single protocol

In the pairs described above, provenance conflates city, study and protocol, which are not separable from one another. One cohort allows them to be dissociated: the Turkish study recruited in two cities under a single protocol, with the same extraction kits and the same diagnostic criteria, and with no difference in sequencing depth between centers (p = 0.78) [16].

Control subjects from Istanbul and Kayseri were separable with an AUC of 0.947, and subjects with Alzheimer’s disease from both cities with an AUC of 0.922. Internal performance was 0.633 at both centers. A model trained in Istanbul reached 0.562 when applied in Kayseri, and the reverse 0.515. The correlation between their importance rankings was 0.003 (p = 0.97), with three taxa shared among the top twenty.

## Discussion

The results show that the separation between cases and controls observed within a cohort is not preserved when the model is applied to another population. The magnitude of that loss is unrelated to geographic distance: in Alzheimer’s disease, transfer between cities of the same country was equivalent to transfer between continents.

The case hardest to attribute to technical variation is the Turkish cohort. Two recruitment centers some seven hundred kilometers apart, with a single extraction and sequencing protocol and no difference in depth between them, produced control populations separable with an AUC of 0.947 and models that do not transfer to each other. That differences exist between distant populations is an accepted observation in the field; that they appear between two cities of the same country, within the same study, places the scale of heterogeneity below the national boundary. This phenomenon converges with what was observed in a metabolic disease cohort within a Chinese province [14], despite differences in condition, population and geographic scale.

### Origin predominates over condition

Comparing the three separability measures puts the phenomenon in perspective. Distinguishing cases from controls within a cohort reached a mean AUC of 0.67 to 0.75 depending on the condition. Distinguishing the cohort of origin of two subjects reached values of 0.94 to 1.00, and did so equivalently using only controls or only subjects with the condition.

The condition does not reduce separability between populations. If the pathology converged toward a common altered state, affected subjects from different populations would be expected to be less distinguishable from one another than controls. No such effect was observed in any of the three conditions. This observation constrains the interpretation of the differential taxa reported in the literature: variation associated with sample provenance is of greater magnitude than that associated with the condition under study.

### Computational harmonization does not resolve comparability

The ten cohorts were processed with the same version of the variant inference algorithm, the same taxonomic reference database and a fixed seed. Despite this, they were separable from one another with AUC values between 0.93 and 1.00.

Differences that precede sequencing DNA extraction protocol, library preparation, sequencer chemistry, depth, recruitment criteria are not corrected by downstream bioinformatic processing [15]. In two of the cohorts, a systematic difference in depth between cases and controls was additionally detected, in opposite directions. The practical consequence is that aggregating cohorts from independent studies requires explicit verification of separability and, where appropriate, batch effect correction [19,20], before interpreting differences between groups as attributable to the condition under study.

The effect of processing decisions on the result is of the same magnitude as that attributed to the condition under study. In one of the autism spectrum disorder cohorts, processing reads in paired-end mode instead of single-end changed the internal AUC by 0.108 points, a value greater than the mean difference between the internal performance of the Parkinson’s disease cohorts and that of the Alzheimer’s disease cohorts. Comparability between studies therefore requires verifying not only data provenance but also the processing decisions applied to each dataset.

### Effect direction and predictive structure are distinct properties

Parkinson’s disease offers the clearest contrast. Eight of 173 genera maintained concordant direction and significance between Finland and Romania, the highest directional concordance of the three conditions; nevertheless, the model trained in Finland performed at chance level in Romania and the correlation between importance rankings was 0.014.

That a taxon is altered in the same direction in two populations does not imply that a classifier using it is transferable. Effect direction is a marginal property of each taxon; predictive structure depends on the joint relationships among all of them, which may differ even when individual directions coincide. This distinction is relevant when interpreting the replication of findings across studies: agreement in the direction of a differential taxon does not constitute validation of a model derived from it.

It should be noted, however, that conclusions concerning Parkinson’s disease rest on a single pair of cohorts, with sample sizes of 133 and 39 subjects, and that this pair additionally spans two different countries. Unlike the Alzheimer’s disease analysis, supported by fifteen cohort pairs of which eight correspond to cities within the same country, the design available for Parkinson’s disease does not permit inference of the within-country pattern in that condition.

### The functional clue

**Most of the** taxa concordant across regions belong to a single functional group. In Alzheimer’s disease, *Lachnospira* and *Faecalibacterium*; in Parkinson’s disease, *Fusicatenibacter*, *Blautia*, *Roseburia*, *Agathobaculum* and *Agathobacter*. All of **these** participate in the production of short-chain fatty acids.

This suggests that functional capacity might be conserved across populations more often than the taxonomic composition that sustains it. Alternative interpretations exist: that the relevant pathophysiological pathways differ between populations, or that part of the discordance is technical in origin. Testing this hypothesis requires validated functional inference and, ultimately, direct measurement of metabolites, available in only one of the cohorts analyzed [12].

### Limitations

This work evaluates separability between groups and its transferability between populations. It does not evaluate diagnostic capacity, nor does it propose criteria for clinical classification.

In cohort pairs drawn from independent studies, provenance conflates city, study and protocol, which are not separable. Only the Turkish cohort allows them to be dissociated by design. No public cohort records comparable dietary data, so the hypothesis of dietary mediation cannot be tested with this material.

Sample support also differs across conditions. The Alzheimer’s disease analysis has fifteen cohort pairs and six cities; autism spectrum disorder, three pairs; and Parkinson’s disease, a single international pair. Statements about the scale at which heterogeneity operates are supported principally by the first.

The Alzheimer’s disease and Parkinson’s disease cohorts use the V3-V4 region and the autism spectrum disorder cohorts the V4 region; magnitudes are not comparable across conditions. The cohorts were moreover designed to answer questions specific to each study, and relevant variables such as gastrointestinal comorbidity, medication and diet are reported in some and not in others, which prevents homogeneous stratification. Gastrointestinal comorbidity, frequent in autism spectrum disorder and a known source of variation in microbial composition [10], is reported with a per-subject score in only one of the three cohorts of that condition.

The decision to process the autism spectrum disorder cohorts in single-end mode, imposed by the deposit format of one of them, reduces the resolution available in the other two. The corresponding sensitivity analysis is presented in the supplementary material.

Three cohorts did not reach significance in the permutation test, and two have reduced sample sizes (n = 35 and n = 39); one of these additionally shows an age difference of 7.3 years between groups, which cannot be adjusted for at that sample size. Finally, analyses by number of features used selection prior to cross-validation, which introduces optimistic bias in those specific estimates.

### Implications

The results suggest that deriving microbiome signatures from individual cohorts requires validation in independent populations before their generality can be assumed, and that such validation cannot be replaced by aggregating findings that agree in direction.

Resolving the underlying question what proportion of the observed heterogeneity is population-related and what proportion technical requires cohorts collected under a single protocol in different populations, with homogeneous clinical phenotyping and direct measurement of metabolites. Available public data do not permit that separation except in exceptional cases of multicenter design, such as the one analyzed here.

## Methods

### Scope of the study

This work evaluates separability between cases and controls in 16S sequencing cohorts and its transferability between populations. It does not evaluate diagnostic capacity, nor does it propose criteria for clinical classification.

### Pre-specified hypotheses

Before running any model, two predictions were stated in writing. The first was that the performance published for the Turkish cohort (AUC 0.63) would be explained by having combined two different recruitment centers, and that analyzing them separately would yield higher performance in each. The second was that the taxa most important to the model would not be the same in different populations, even if the direction of their abundance did coincide.

The first prediction was not confirmed: analyzed separately, both centers reached an AUC of 0.633, the same value as the combined set. Heterogeneity between cities did not manifest where it was expected as a loss of performance within each center — but in the transfer between them and in the discordance between their discriminant taxa. The second prediction was confirmed in all three conditions.

### Search and admission criteria

The European Nucleotide Archive, the Sequence Read Archive, QIITA and Zenodo were searched using terms relating to the three conditions combined with “microbiome”, “gut microbiota” and “16S”. The search was complemented with the data availability statements of publications in the field. An admission protocol was applied to each identified deposit, verified against public metadata before downloading sequence files: sample type restricted to feces; 16S amplicon strategy, excluding shotgun metagenomics and RNA-Seq; hypervariable region matching the other cohorts of the same condition; Illumina platform; one sample per subject, or the possibility of selecting one; group labels accessible in public metadata; institution and location of origin confirmed in the deposit metadata; and absence of intervention prior to sampling, or an isolable baseline.

The hypervariable region was verified by inspecting the first bases of the reads in a representative file per deposit. This criterion excluded cohorts widely cited in the field, among them those of Scheperjans [8] (V1-V3 region, 454 platform) and Kang [10] (V2/V3 region, pyrosequencing), as not comparable with the other cohorts of their respective conditions.

### Sequence processing

Sequences were processed with DADA2 version 1.34 in R 4.4.2, with the seed fixed at 42.

Where primers were present, they were removed by sequence localization using vmatchPattern from the Biostrings package, allowing degenerate bases and up to two mismatches; only the read pair in which both ends contained the corresponding primer was retained. Fixed-position trimming was not used, given that several cohorts contain heterogeneity spacers of variable length.

Truncation parameters were calibrated per cohort according to the actual read length after trimming. maxN = 0, maxEE = c(2,5), truncQ = 2 and PhiX removal were applied. The error model was learned independently for forward and reverse reads; pair merging used a minimum overlap of twelve bases and chimeras were removed by the consensus method. Taxonomic assignment was performed against SILVA nr99 version 138.2, with search in both orientations; sequence variants were aggregated at genus level and those unassigned at that rank were discarded.

The three autism spectrum disorder cohorts were processed in single-end mode on the forward read truncated to 145 bases. One of them is deposited only as a forward read and therefore does not permit pair merging; processing the other two in paired-end mode would have introduced a processing difference between cohorts of the same condition.

The effect of this decision was evaluated by reprocessing in paired-end mode the two cohorts that allow it. Internal performance increased in one of them (from 0.595 to 0.703; p changed from 0.163 to 0.003) and was maintained in the other (0.691 to 0.694), while transfer between the two declined from 0.530 and 0.524 to 0.447 and 0.418, and between-cohort separability remained unchanged (0.998 and 0.999). Single-end processing therefore yields higher transfer estimates than paired-end processing, and so constitutes the conservative choice with respect to the conclusion of this work (Table S3).

In the Finnish cohort, of longitudinal design [17], a single sample per subject was retained, the earliest by collection date. In the Italian cohort, only baseline samples were used, prior to the probiotic intervention. In the Shanghai cohort, which additionally included a mild cognitive impairment group [5], only the Alzheimer’s disease and control groups were used.

### Data preparation and classification models

Genus-level count matrices were converted to relative abundance and transformed by centered log-ratio, using a pseudocount of 1×10⁻⁶ for zeros. Cross-cohort analyses used the genera common to all cohorts of the condition, filtered by a minimum prevalence of 20% in each of them.

The pseudocount value was set at 1×10⁻⁶, below the lowest detectable relative abundance given the sequencing depths of the set. A sensitivity analysis over the range 1×10⁻⁷ to 1×10⁻⁴ produced mean variations of 0.025 AUC points, without altering the significance of any cohort (Table S1).

Random forests were used with 500 trees, max_features = sqrt and balanced class weighting. Internal separability was estimated by stratified ten-fold cross-validation, repeated twenty times with different seeds, and the mean is reported. Confidence intervals were obtained by bootstrap with 1,000 resamples of the out-of-fold predictions, and the null hypothesis was tested by permutation test with 300 permutations of the labels. Transfer was evaluated by training the model on the whole of one cohort and applying it to the whole of another, restricted to shared features. Between-cohort separability was estimated by training a classifier to predict cohort of origin, disregarding clinical group, separately on controls and on subjects with the condition.

All performance evaluation was carried out on out-of-sample predictions; values obtained on training data are not reported.

### Feature importance and effect size

SHAP values were calculated with TreeExplainer over twenty models fitted with independent seeds, averaging the absolute value per feature. Within-cohort stability was evaluated by Spearman correlation between all pairs of rankings obtained with different seeds; between-cohort concordance, by Spearman correlation between the averaged rankings and by the overlap among the twenty most important features.

Effect size between groups was quantified by Cliff’s delta on relative abundances, with confidence intervals from 500 bootstrap resamples, and significance by the Mann-Whitney U test. For the regional contrast, the cohorts of each condition were grouped by region and the effect was calculated separately in each group. A genus was considered concordant if it had the same sign in both regions, magnitude equal to or greater than 0.10 in both, and significance in both; a confirmed reversal was defined as a genus with opposite sign, magnitude equal to or greater than 0.10 in both regions, and non-overlapping confidence intervals.

## Data Availability

All cohorts analyzed are publicly accessible. The sequence data are deposited in the European Nucleotide Archive (PRJEB27564, PRJEB89799) and in the NCBI Sequence Read Archive (PRJNA315645, PRJNA489760, PRJNA533120, PRJNA554111, PRJNA633959, PRJNA734525, PRJNA953169, PRJNA1046003). The processing and analysis code, with the documentation required to reproduce the results, is available at https://github.com/jtellolatam/microbiome-radar-geographic-heterogeneity and permanently archived in Zenodo (https://doi.org/10.5281/zenodo.24374407).

https://www.ebi.ac.uk/ena/browser/home

https://www.ncbi.nlm.nih.gov/bioproject/

https://github.com/jtellolatam/microbiome-radar-geographic-heterogeneity

https://doi.org/10.5281/zenodo.24374407

## Data and code availability

All cohorts analyzed are publicly accessible. The sequence data are deposited in the European Nucleotide Archive (PRJEB27564, PRJEB89799) and in the NCBI Sequence Read Archive (PRJNA315645, PRJNA489760, PRJNA533120, PRJNA554111, PRJNA633959, PRJNA734525, PRJNA953169, PRJNA1046003). The processing and analysis code, with the documentation required to reproduce the results, is available at https://github.com/jtellolatam/microbiome-radar-geographic-heterogeneity and permanently archived in Zenodo (https://doi.org/10.5281/zenodo.24374407). The supplementary material includes Tables S1 and S2, with the pseudocount sensitivity analysis, and Table S3, with the sensitivity analysis of processing mode in the autism spectrum disorder cohorts.

## Declarations

### Author contributions

Johann Tello Meryk conceived the study, designed the cohort admission protocol, performed the bioinformatic processing and statistical analysis, and wrote the manuscript.

### Ethical considerations

The author confirms that this study did not require ethical approval. All data analyzed are publicly available, de-identified 16S rRNA gene sequencing datasets, previously collected under the ethical approvals reported by the original studies and deposited in public repositories. No new participants were recruited, and no identifiable data were accessed.

## Supplementary material

**Table S1.** Pseudocount sensitivity analysis. Internal AUC of each cohort, obtained by stratified ten-fold cross-validation repeated twenty times, using different pseudocount values in the zero substitution prior to the CLR transformation. Values are on the scale of relative abundances.

| **Cohort** | **Condition** | **1×10<sup>−7</sup>** | **1×10<sup>−6</sup>** | **1×10<sup>−5</sup>** | **1×10<sup>−4</sup>** | **Range** |
|---|---|---|---|---|---|---|
| Shanghai | Alzheimer’s | 0.978 | 0.983 | 0.985 | 0.989 | 0.010 |
| Lishui | Alzheimer’s | 0.856 | 0.862 | 0.862 | 0.860 | 0.006 |
| Chongqing | Alzheimer’s | 0.712 | 0.710 | 0.675 | 0.671 | 0.041 |
| Istanbul | Alzheimer’s | 0.656 | 0.657 | 0.683 | 0.687 | 0.031 |
| Kayseri | Alzheimer’s | 0.629 | 0.621 | 0.617 | 0.619 | 0.011 |
| Brescia | Alzheimer’s | 0.686 | 0.683 | 0.703 | 0.699 | 0.020 |
| Nanjing | Autism spectrum | 0.959 | 0.959 | 0.957 | 0.950 | 0.009 |
| Xiamen | Autism spectrum | 0.591 | 0.595 | 0.622 | 0.627 | 0.036 |
| Pasadena | Autism spectrum | 0.707 | 0.691 | 0.693 | 0.706 | 0.015 |
| Helsinki | Parkinson’s | 0.674 | 0.658 | 0.660 | 0.645 | 0.029 |
| Iași | Parkinson’s | 0.729 | 0.755 | 0.794 | 0.790 | 0.065 |
| Mean |  | 0.743 | 0.743 | 0.750 | 0.749 | 0.025 |

Range: difference between the maximum and minimum value of each row. No cohort changes its statistical significance within this interval.

**Table S2.**
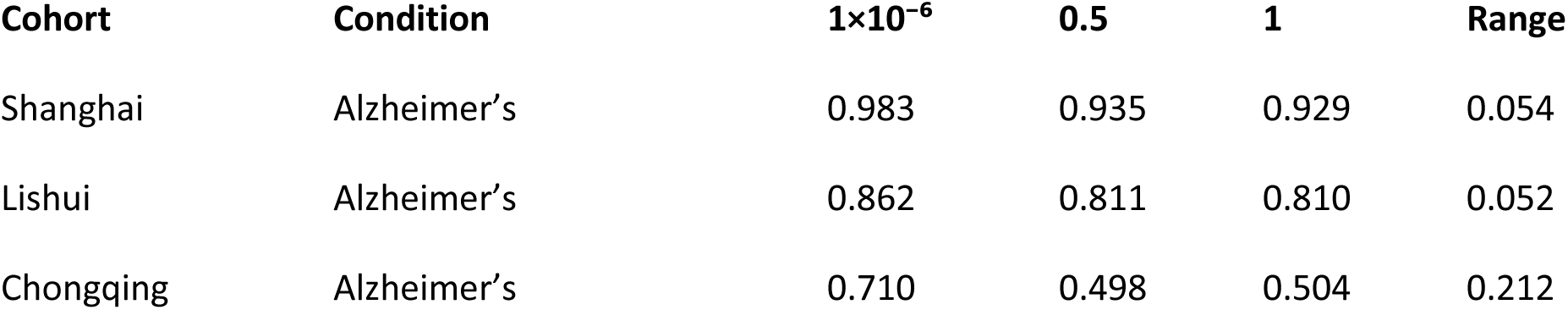
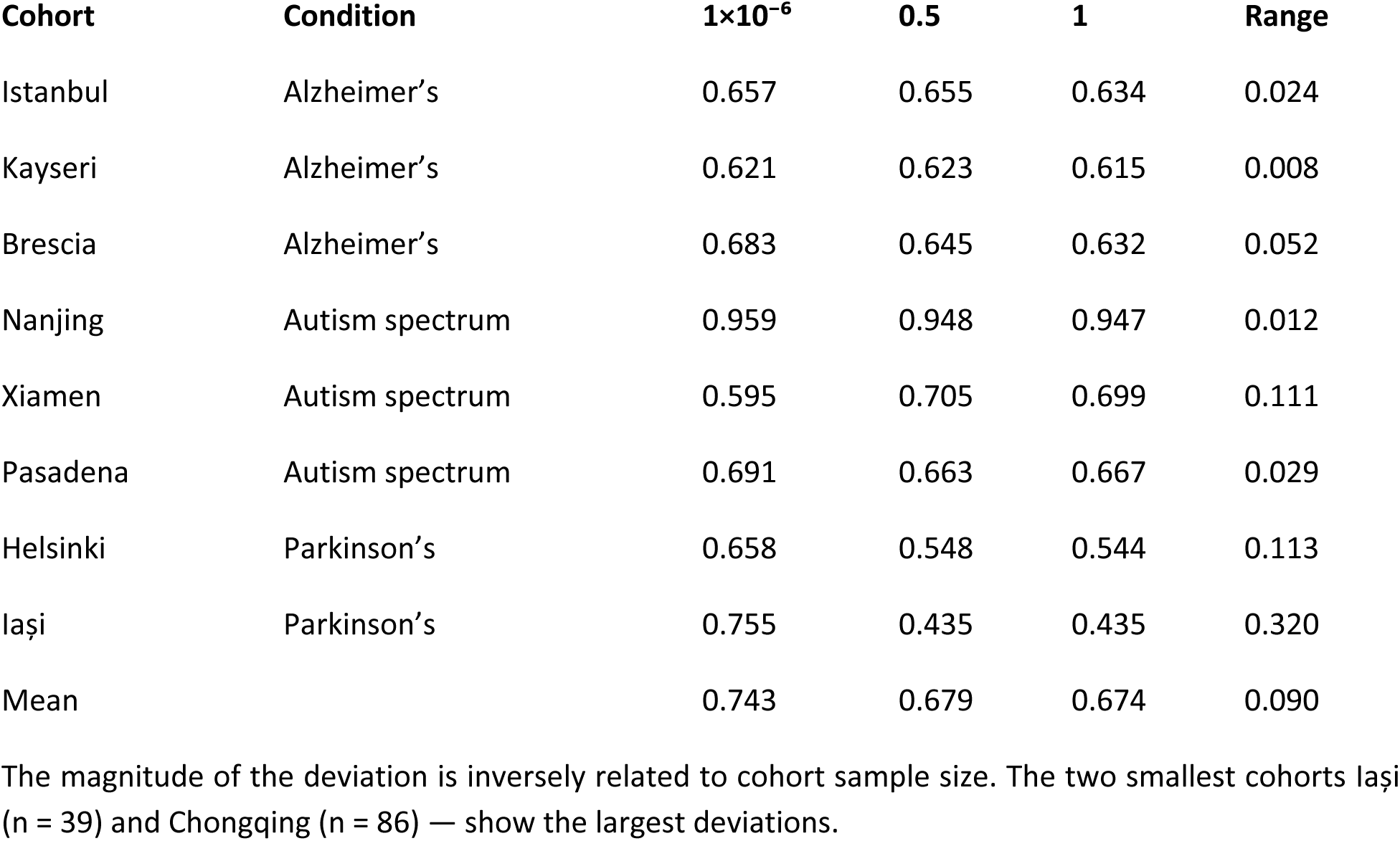
Sensitivity to pseudocount values of a different scale. The values 0.5 and 1 are customary when substitution is applied to raw counts.

| **Cohort** | **Condition** | **1×10<sup>−6</sup>** | **0.5** | **1** | **Range** |
|---|---|---|---|---|---|
| Shanghai | Alzheimer’s | 0.983 | 0.935 | 0.929 | 0.054 |
| Lishui | Alzheimer’s | 0.862 | 0.811 | 0.810 | 0.052 |
| Chongqing | Alzheimer’s | 0.710 | 0.498 | 0.504 | 0.212 |
| **Cohort** | **Condition** | **1×10<sup>−6</sup>** | **0.5** | **1** | **Range** |
| Istanbul | Alzheimer’s | 0.657 | 0.655 | 0.634 | 0.024 |
| Kayseri | Alzheimer’s | 0.621 | 0.623 | 0.615 | 0.008 |
| Brescia | Alzheimer’s | 0.683 | 0.645 | 0.632 | 0.052 |
| Nanjing | Autism spectrum | 0.959 | 0.948 | 0.947 | 0.012 |
| Xiamen | Autism spectrum | 0.595 | 0.705 | 0.699 | 0.111 |
| Pasadena | Autism spectrum | 0.691 | 0.663 | 0.667 | 0.029 |
| Helsinki | Parkinson’s | 0.658 | 0.548 | 0.544 | 0.113 |
| Iași | Parkinson’s | 0.755 | 0.435 | 0.435 | 0.320 |
| Mean |  | 0.743 | 0.679 | 0.674 | 0.090 |

The magnitude of the deviation is inversely related to cohort sample size. The two smallest cohorts Iași (n = 39) and Chongqing (n = 86) — show the largest deviations.

**Table S3.** Sensitivity analysis of processing mode in the autism spectrum disorder cohorts. The two cohorts deposited with paired reads were reprocessed in paired-end mode (truncLen 145/140) and compared with the single-end processing used in the main analysis. The third cohort of this arm is deposited only as a forward read and does not permit this comparison.

| Measure | Single-end | Paired-end | Difference |
|---|---|---|---|
| Internal AUC · Xiamen | 0.595 | 0.703 | +0.108 |
| 95% CI | [0.474 – 0.723] | [0.572 – 0.800] |  |
| p (permutation) | 0.163 | 0.003 |  |
| Internal AUC · Pasadena | 0.691 | 0.694 | +0.003 |
| 95% CI | [0.551 – 0.794] | [0.578 – 0.807] |  |
| p (permutation) | 0.010 | 0.007 |  |
| Transfer Xiamen → Pasadena | 0.530 | 0.447 | −0.083 |
| Transfer Pasadena → Xiamen | 0.524 | 0.418 | −0.106 |
| Cohort separability | 0.998 | 0.999 | +0.001 |
| Separability · controls only | 0.990 | 0.991 | +0.001 |
| Shared genera | 82 | 73 |  |

Paired-end processing increases internal performance and reduces transfer between cohorts. Between-cohort separability remains unchanged. The single-end mode used in the main analysis therefore yields higher transfer estimates, and constitutes the conservative option with respect to the conclusion of this work.

